# Synthetic Data to Lower Barriers Towards Equitable Artificial Intelligence in Rapid Diagnostic Test Interpretation

**DOI:** 10.1101/2025.02.25.25322677

**Authors:** Elliott Rogers, Valérian Turbé, Dickman Gareta, Carina Herbst, Rachel A. McKendry, Kobus Herbst, Maryam Shahmanesh

**Affiliations:** London Centre for Nanotechnology and Division of Medicine, University College London, London, UK; Institute of Global Health, University College London, London, UK; Africa Health Research Institute, Durban, South Africa; Institute of Social and Preventive Medicine, University of Bern, Bern, Switzerland; Graduate School for Health Sciences, University of Bern, Bern, Switzerland; School of Laboratory Medicine and Medical Sciences, University of KwaZulu Natal, Durban, South Africa; DSI-SAMRC South Africa Population Research Infrastructure Network, Durban, South Africa

## Abstract

Rapid diagnostic tests (RDTs) support affordable disease diagnosis. Machine learning (ML) can improve RDT interpretation but often relies on large, proprietary, and costly real-world image libraries. We present SynSight – a ML-enabled RDT segmentation and classification pipeline trained on synthetic data. Validated on HIV (98% sensitivity, 99% specificity) and COVID-19 RDTs (up to 99% accuracy), SynSight enables rapid ML training without real-world training images, keeping pace with new RDT development.

## Main text

### Rapid diagnostic tests: challenges and machine learning opportunities

Lateral flow tests are indispensable rapid diagnostic tests (RDT) valued for delivering results within minutes and meeting the REASSURED criteria^1^. Simple RDTs, such as COVID-19 RDTs, display a red test line to indicate antigen detection, while multiplex RDTs detect multiple targets simultaneously, such as Immunoglobulin G (IgG) and IgM antibodies. RDT use for managing acute and chronic diseases is ever increasing^1^. Over 3 billion COVID-19 RDTs were performed by late-2022^2^. In 2024, GAVI initiated the largest global deployment of 1.2 million cholera RDTs^3^. HIV RDT use is projected to reach 206 million by 2027^4^. However, RDT performance – reported through sensitivity, specificity, and accuracy – can be undermined by subjective human interpretation, leading to result misclassifications^5–7^.

Machine learning (ML) supports interpretation of RDT results^8^, including for HIV^7,9^, COVID-19^9–13^, influenza^14^, and malaria^14–17^, and can be deployed via increasingly available mobile devices^8,18^. However, training ML models often relies on large, proprietary real-world image libraries or pre-existing models. These are typically clinically sensitive, difficult to access, and expensive, with privacy and data-sharing restrictions introducing delays, costs, and barriers to access^7,11,12,19^. This can disadvantage implementers in low- or middle-income countries (LMICs), where data capture and access can be constrained by cost and infrastructure, hindering the development and deployment of convolutional neural networks (CNNs).

In 2021, our previous study (Turbé et al.) introduced ClassifyAI, a CNN trained on over 6,000 operationally acquired HIV RDT images collected by sixty trained workers over two years which were manually cropped to the region of interest (ROI) and labelled^7^. In 2023, a state-of-the-art few-shot learning model, AutoAdaptPOC, reduced the number of real-world training images to 1/50^th^ of those previously needed; however, it still relied on a base model trained on hundreds of real-world images^9^.

### SynSight: synthetic data for training RDT machine learning models

Synthetic data offers a promising alternative to real-world data and is used in healthcare to address data paucity, class imbalance, model generalisability, cost and privacy^20,21^. Broadly, synthetic images can be created by either training a model on real-world data to create similar images (e.g. Variational Autoencoders (VAEs), Generative Adversarial Networks (GANs)), with accompanying privacy concerns^21^, or by using virtual environments (e.g. mechanistic simulators) to generate labelled datasets using no real-world data^22^. Generative models are a powerful image generation method but can be challenging to control and resulting models can collapse when trained on recursively generated data^23,24^. We instead used a mechanistic simulator to allow the control of all features of the virtual environment, such as lighting (e.g. intensity) and camera parameters (e.g. angle) (Supplementary Figure 1). Mechanistic simulators have been highlighted for their potential in computer vision for diagnostics^22^. A systematic comparison of generative versus mechanistic data generation and of the resulting ML models was beyond the scope of this study, as the difficulty of controlling label accuracy in generative outputs prevented a fair comparison.

Herein, we present SynSight, a two-stage ML-enabled RDT processing pipeline consisting of ROI segmentation models (Fig. 1a) and result classification models (Fig. 1b) trained on mechanistically generated synthetic data and publicly available models (MobileNetV2^25^ and You-Only-Look-Once (YOLO) v11^26^). A single real-world image can be processed and classified using models trained on synthetic data (Fig. 1c). We used the “train on synthetic, test on real” (TSTR) versus “train on real, test on real” (TRTR) evaluative framework^22,27^ to compare the performance of published models trained on hundreds to thousands of real-world images (TRTR) to our models trained on synthetic data (TSTR). This framework evaluates a model’s generalisability to real-world settings, a key challenge of synthetic data. We validated SynSight on four RDTs, two disease targets (HIV and COVID-19) and both antigen and antibody RDTs (Fig. 2 and 3). SynSight marks the next step in training RDT ML models, reducing real-world training images from thousands (ClassifyAI^7^), to hundreds (AutoAdaptPOC^9^), to none (SynSight, this work).

**Fig. 1:**
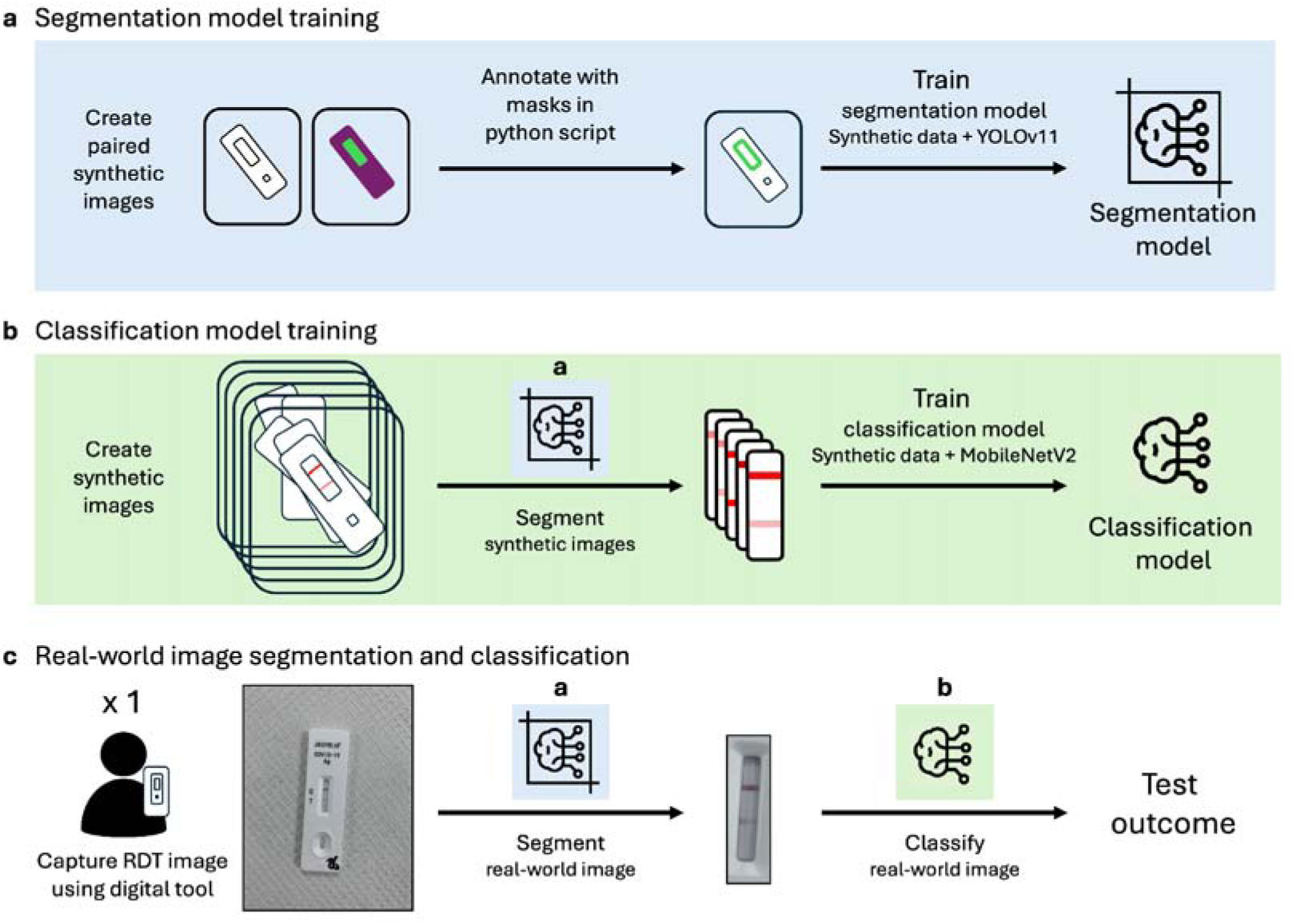
Schematic of (a) segmentation and (b) classification model training on synthetic data and (c) processing a real-world image. a) Segmentation model training is facilitated by automating the ROI annotation of a synthetic image library by using python scripts implementing masks and paired synthetic images. The resulting annotated synthetic image library is used with You-Only-Look-Once v11 (YOLOv11) state-of-the-art segmentation model to train a ROI segmentation model. b) Synthetic images of full RDTs are segmented using the model trained in (a). Class balanced datasets of varying results can be created, including for strong positives, weak positives, negatives, invalid results, and results underrepresented in real-world image libraries. These images can be used with MobileNetV2 to train classification models. c) Single real-world RDT results (real-world image collected by Arumugam et al.) can be classified by implementing the (a) segmentation model and (b) classification model trained on synthetic data. This allows image segmentation and classification for RDTs with no existing large real-world image libraries.

**Fig. 2.**
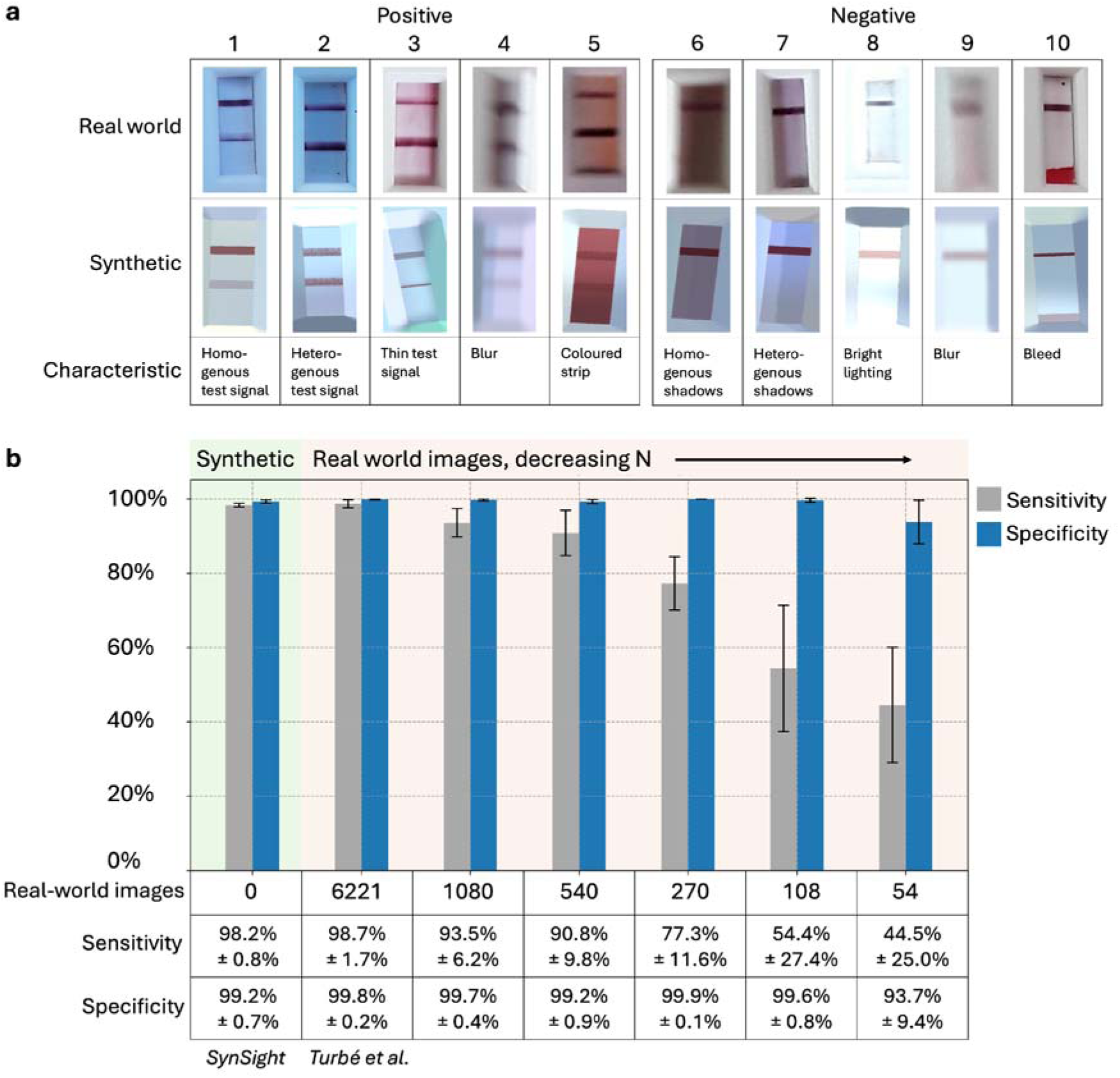
Comparison of (a) real-world HIV RDT images and synthetic images, and (b) models trained on synthetic images (SynSight, this work), 6,221 real-world images (Turbé et al., 2021) and decreasing real-world image training libraries. a) Synthetic images generated in 3D modelling software Unity showing varying test result (positive (1-5) and negative (6-10)), line intensity (homogenous (1), heterogenous (2), and thin (3)), blur (4 and 9), background strip colour (5), shadows (homogenous (6) and heterogenous (7)), lighting (dark (6) and bright (8)) and bleed (10), compared to real-world HIV RDTs (Turbé et al). b) Plot showing the average sensitivity and specificity of HIV RDT classification using our classification model trained on synthetic data (TSTR, synthetic training n=8,000, real-world training n=0, test n = 5,685; green shading) compared to our previous model trained on thousands of real-world images (TRTR, real-world training n=6,221)^7^ and decreasing number of real-world images (TRTR, real-world training n=1080 to n=54; orange shading). The threshold for the synthetic model was calculated using 18% of the real-world dataset, as per our previous publication^7^. The TRTR model was trained using 10-fold cross-validation, with sensitivity and specificity obtained using a hold-out dataset (TRTR, test n=693)^7^. Error bars and ± represent standard deviation from the mean.

To start, we mechanistically generated diverse synthetic training image libraries in minutes, significantly faster than the weeks to years required to collect real-world data (Table 1)^7,10^. Next, we trained RDT ROI segmentation models, a precursor to ML classification^7,9,28^. Mechanistic image generation was leveraged to create paired images which were automatically annotated using python scripts (Fig. 1a). 99.9% (1093/1094) of real-world COVID-19 RDTs produced a segmented output. Localisation accuracy compared to manual annotation was high across all three RDTs (mean IoU: DeepBlue 0.86, 95% CI 0.84-0.87; Jinwofu 0.83, 95%CI 0.80-0.87; EcoTest 0.83, 95%CI 0.81-0.85), evidencing sim-to-real generalisation (Supplementary Figure 2, Supplementary Table 1). We also developed an augmented-reality RDT segmentation model which could guide smartphone data capture (Supplementary Figure 3).

**Table 1.** Comparison of machine learning (ML) model architecture and real-world training images used for three models: ClassifyAI (our previous work), AutoAdaptPOC (2023), and SynSight (this work). ClassifyAI (2021), trained on data collected over years, features manual segmentation of regions of interest (ROI) and a classification model built on MobileNetV2 fine-tuned on 5,528 real-world images. AutoAdaptPOC (2023), trained on real-world data collected over weeks, features automated segmentation using a Mask R-CNN, an instance segmentation model, pre-trained on 60 real-world images and 20-75 images to adapt the model to each new RDT. AutoAdaptPOC classifies images using a pretrained ResNet18 model trained on 383 real-world images and adapted using supervised contrastive learning on 10-14 labelled images. SynSight is a ML-enabled RDT segmentation and classification pipeline trained on synthetic data generated in minutes. Paired synthetic images are generated and automatically annotated using python scripts to produce a labelled ROI dataset. The annotated synthetic image library is then used to train the pre-trained You-Only-Look-Once (YOLO) v11 nano segmentation model, a state-of-the-art open-source segmentation model. SynSight classifies images using a classification model built on MobileNetV2 pretrained on ImageNet and fine-tuned on synthetic images and no real-world training images.

|  |  | 2021<br>ClassifyAI (previous<br>work) | 2023<br>AutoAdaptPOC | 2025<br>SynSight (this work) |
| --- | --- | --- | --- | --- |
| Time to model |  | Years | Weeks - months | Minutes - days |
| Segmentation | Model architecture | Manual | Mask R-CNN | YOLOv11 |
|  | Real-world training<br>images |  | 60 (pretrain),<br>20-75 per new kit | 0 |
| Classification | Model architecture | MobileNetV2 | ResNet-18 +<br>supervised contrastive<br>few-shot learning | MobileNetV2 |
|  | Real-world training<br>images | 5,528 | 383 - 397 | 0 |

For the classification model, synthetic image generation was informed by our previous studies and literature^6,7,10^. Synthetic libraries can be created to improve model performance on challenging images by increasing the frequency of weak positives^10^ (Fig 2a. 1 and 4), underrepresented edge cases (e.g. blood in the ROI^10^, Fig 2a. 10), and rare images (e.g. no control line^7^). Synthetic libraries allow controlled class distributions, unlike real-world libraries which can suffer from class-imbalance (e.g. 1:8 positive to negative ratio for ClassifyAI^7^).

For the HIV RDT, we trained a classification model on 8,000 synthetic images and achieved a TSTR performance of 98.2% (±0.8%) sensitivity, 99.2% (±0.7%) specificity, and 99.2% (±0.6%) accuracy (± standard deviation across 10 random subsets; test n=5,685). This was comparable to our previous model, ClassifyAI, trained on 6,221 real-world images (TRTR 98.7%±1.7% sensitivity, 99.8%±0.2% specificity, accuracy not reported, test n=693)^7^ (Fig. 2). We also evaluated SynSight against ClassifyAI when trained on fewer real-world images. ClassifyAI’s sensitivity dropped to 77.3% (±11.6%), over 20 percentage points lower than SynSight (Fig. 2b), when trained on 270 real-world images. This highlights the strength of the synthetic approach when limited real-world libraries are available.

Next, we applied the full SynSight pipeline, including a single segmentation and a single classification model applied across all RDTs, to two antigen and one antibody COVID-19 RDT. Training and threshold selection only used synthetic images, allowing classification of complete image libraries. Classification accuracy of two antigen RDTs was 99.3% (95%CI 97.5-99.9, test n=283; TRTR:98.9%, test n=184; DeepBlue) and 97.0% (95%CI 93.6-98.9, test n=200; TRTR:100%, test n=104; Jinwofu), similar to the performance of the few-shot learning approach trained on approximately 400 real-world images^9^. For the antibody RDT, the base-model training set used in the few-shot learning approach, SynSight achieved 96.6% accuracy (95%CI 94.8-97.9, test n= 610; TRTR:100%, test n=102; EcoTest) (Figure 3)^9^.

### Evaluating real-world images and labels

Real-world images and provided ground truth labels remain important to validate ML models. Consistent with prior work, HIV classification thresholds were calculated using a subset of real-world data, while COVID-19 thresholds were calculated using synthetic data. All performance evaluation was conducted on real-world images using the provided ground truth labels (Supplementary Table 2). However, real-world images and labels can have issues, and disagreements between ML and human interpretation should therefore be visually evaluated. False negatives (FN - ML negative, ground truth positive) can be difficult to verify as weak test lines may not be captured in images, a limitation of ML RDT analysis. False positives (FP - ML positive, ground truth negative) are easier to verify as a visible test line indicates incorrect ground truth labelling. We examined FP and FN results to evaluate ground truth label accuracy and to explore SynSight’s performance across challenging image conditions (Supplementary Table 3).

SynSight outperformed the benchmark model on the DeepBlue RDT but underperformed on the Jinwofu RDT. A comparison of SynSight’s Jinwofu FP results (n=4) (Fig. 3c J2-5) alongside a true positive (J1) and negative (J6) revealed a visible weak test line in three of the processed ROI results (Fig. 3c J2-4), suggesting the provided ground truth labels are incorrect. Further, SynSight incorrectly classified 1/283 DeepBlue RDTs with excessive streaking (where nanoparticles are deposited throughout the ROI) and 2/610 EcoTest RDTs with artefacts presenting as marks near test line locations as FP (Supplementary Figure 4). For FN results, SynSight incorrectly classified 19/610 EcoTest RDTs (8 with weak test lines) and 2/200 Jinwofu RDTs (affected by shadows and poor segmentation) as FN (Supplementary Figure 5). This demonstrates how ML-human disagreements can flag labelling errors, supporting label-auditing and adjudication workflows.

**Fig. 3.**
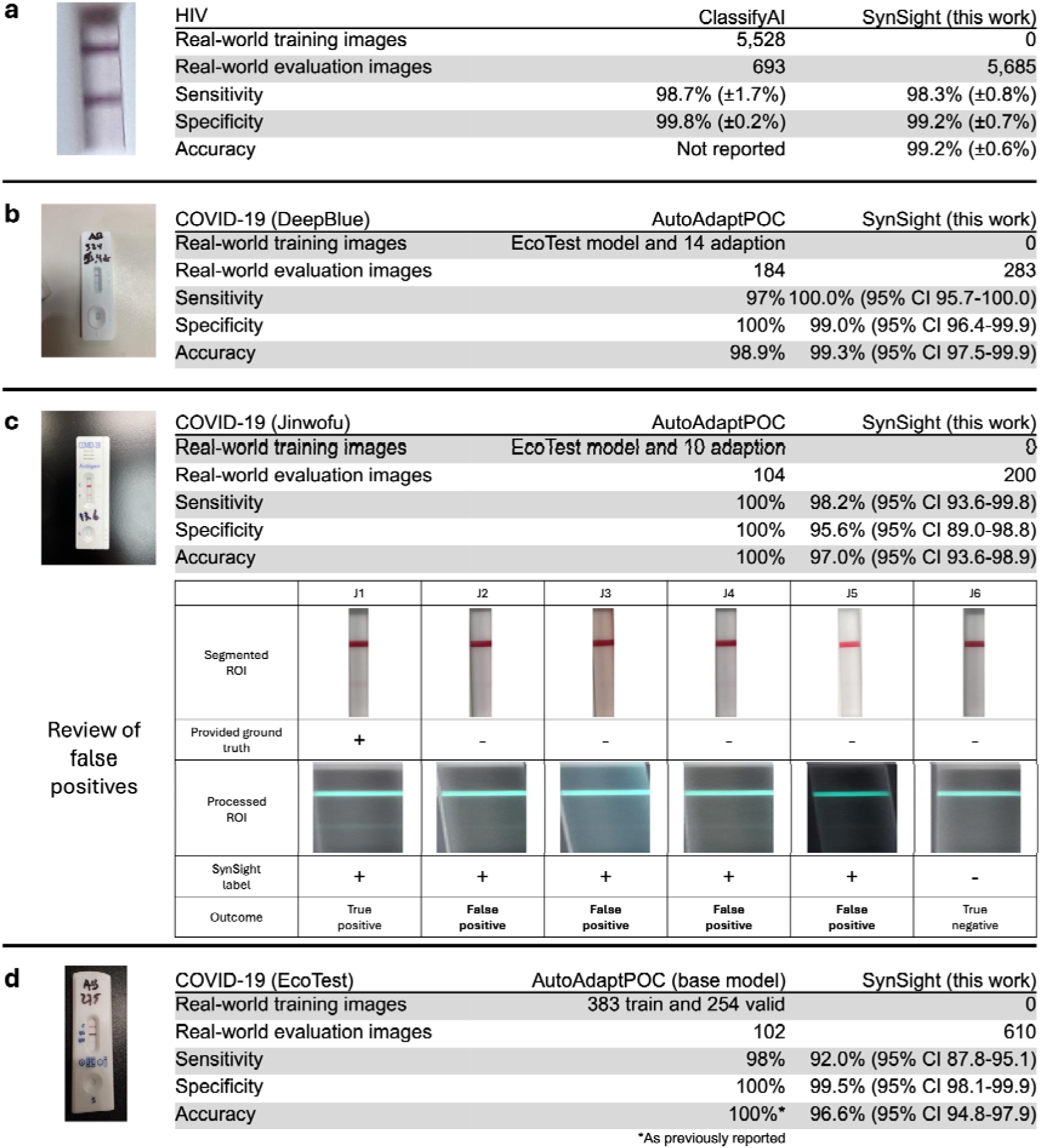
Comparison of the performance of SynSight, our machine learning models trained on synthetic data and no real-world training images, to published model performance of one HIV antibody (rapid diagnostic test) RDT and two COVID-19 antibody and one COVID-19 antigen RDTs. SynSight (0 real-world training images) matches the HIV RDT sensitivity (98.3%) and specificity (99.2%) compared to the benchmark model, ClassifyAI (5,528 real-world training images, 98.7% sensitivity, 99.8% specificity), and for the COVID-19 RDTs, meets the performance on the DeepBlue COVID-19 RDT (SynSight: 100% sensitivity, 99.0% specificity, 99.3% accuracy; comparative AutoAdaptPOC benchmark model: EcoTest base model and 14 adaptation training images, 97.0% sensitivity, 100% specificity, 98.9% accuracy), the Jinwofu RDT (SynSight: 98.1% sensitivity, 95.6% specificity, 97.0% accuracy; AutoAdaptPOC: EcoTest base model and 10 adaptation training images: 100% sensitivity, 100% specificity, 100% accuracy), and the EcoTest RDT (SynSight: 92.0% sensitivity, 99.5% specificity, 96.6% accuracy; AutoAdaptPOC: EcoTest base model (383 training and 254 validation training images), 98% sensitivity, 100% specificity, 100% accuracy). Segmented Regions of Interest (ROIs) and processed ROIs are displayed for all false positives predicted by SynSight (J2-5), alongside a true positive (J1) and true negative (J6), based on provided ground truth labels. Test lines visible in J2-4 suggest the provided ground truth labelling is incorrect. Due to synthetic training, for the HIV RDT, SynSight can evaluate a real-world image library over eight times larger than ClassifyAI, and 1,093 COVID-19 RDTs compared to 390 RDTs of AutoAdaptPOC. HIV metrics are reported alongside standard deviation from the mean, COVID-19 metrics are reported alongside 95% exact binomial (Clopper–Pearson) confidence intervals.

Limitations of our approach include poor generalisability of ML models trained on mechanistically generated synthetic data when parametric assumptions (e.g. camera angle, test line morphology) fail to capture real-world image variability, leading to poor performance, bias, and confabulation – plausible but incorrect predictions based on patterns in synthetic images. Expert-informed synthetic image library design, focusing on known classification challenges, iterative improvement of image generation in response to observed failure modes and feedback, and standardised real-world image capture protocols could improve generalisability of models trained on synthetic data. For example, poor performance on low-lighting image environments could be addressed by generating more low-light examples in the synthetic library. Synthetic data mitigates real-world label noise during training but does not eliminate it, as noise can persist during evaluation, as highlighted by SynSight identifying test lines missed in ground truth labelling.

### Generalisability to medical diagnostics

Medical diagnostic imaging outputs range from simple to complex, with mechanistic simulation applicability varying accordingly. Imaging modalities can be categorised into three environments: directly suitable; possibly suitable, requiring modality-specific simulation; and challenging and speculative.

Directly suitable environments include visually constrained, object-based images, such as RDT result images. Results appear planar, have a regular, predictable format, are captured on similar edge devices (mobile phones) under a fixed range of lighting conditions, with classes distinguished through a simple test line presence/absence. The images are low-dimensional and so suitable for physics-based mechanistic image generation.

Possibly suitable environments which warrant further exploration include chest x-rays for respiratory diseases and fundus photography for ophthalmopathy. These imaging environments remain constrained but have complex outputs with more class-defining features which would require advanced mechanistic simulation and may benefit from hybrid training approaches that combine real-world and synthetic data. Time-series imaging for the same individual imaged at consecutive time points, such as for tuberculosis chest x-ray monitoring, adds a further dimension to synthetic image generation.

Challenging and speculative environments include CT, MRI, ultrasound, and echocardiography. These advanced imaging modes and resulting images are significantly affected by patient variability (anatomy and pathology), modality-specific acquisition physics (probe pressure, angle, and acoustic propagation) and object motion, and produce volumetric (3D) data. Further, echocardiography and cardiac MRI have time-series data for the same patient captured within a single acquisition time point across cardiac cycles, resulting in 4D data. Purely mechanistic simulation of these imaging modes would be challenging and may not prove effective, as 3D and 4D datasets would need to be defined through complex, interacting parameters that accurately represent individuals and individual classes. For these environments, increasingly powerful generative models are likely to prove more effective than mechanistic simulation alone.

### Towards equitable diagnostic AI

SynSight could support more equitable AI development and deployment for low-dimensional diagnostic imaging by reducing the need for data sharing agreements or extensive resources for data capture and model training (conducted on both CPU and GPU). SynSight is trained on synthetic data generated using widely available 3D modelling software (Unity) and on open-source models (MobileNetV2, YOLOv11), instead of closed or proprietary base models. By tailoring the available mechanistic data generation methods to their own target RDT formats, implementers in LMICs could create and own datasets and train and own ML models. The secondary image analysis presented here should be expanded to prospective implementation to evaluate real-world feasibility and usability across multiple image capture devices.

In conclusion, synthetic data and digital tools can support the use of RDTs, improving interpretation by reducing false positives and negatives. SynSight segments and classifies four RDTs, including antigen and multiplex antibody RDTs for two different diseases, with similar performance to models trained on hundreds and thousands of real-world images. SynSight mitigates privacy concerns associated with training on sensitive clinical data and could reduce barriers to developing, accessing, and owning customised training image libraries and training ML models. Synthetically trained ML-enabled digital tools promise scalable, automated, accurate interpretation and quality control of RDTs, supporting the rapid development and deployment of accurate models for new and existing tests.

## Methods

### Ethics

The work presented are secondary analyses of existing real-world image libraries of HIV and COVID-19 RDTs. Ethical approval for the Africa Health Research Institute’s (AHRI) HIV image library was collected as part of the demographic surveillance study, with ethics granted by the Biomedical Research Ethics Committee of the University of KwaZulu-Natal, South Africa (no. BE435/17)^7,29^. The ethics for the collection of the COVID-19 RDT image library are outlined in a previous publication^9^.

### Real-world data

Images of HIV RDTs were collected by the AHRI. Image capture of Advanced Quality One Step Anti-HIV (1&2) Test (InTec Products, Inc.)) tests was embedded into trained staff workflows in Kwazulu-Natal, South Africa, using minimal hardware (Samsung SM-P585 tablet and customised imaging tray), as previously described^7^. The two antigen COVID-19 RDTs were the Anhui DeepBlue SARS-CoV-2 Antigen Test and the Jinwofu SARS-CoV-2 Antigen Rapid Test. The antibody COVID-19 RDT was the AssureTech EcoTest COVID-19 IgG/IgM Antibody Test kit. These COVID-19 RDTs were conducted and imaged by researchers at Columbia University using nasopharyngeal swabs from Mayo Clinic Hospital patients (Mayo Clinic IRB 20-010688)^9^. 25 duplicate RDT images and 1 test labelled ‘handmarked’, with a test line clearly marked on the RDT, were identified for the EcoTest RDT and excluded from analyses. We have developed open-source hardware to facilitate the capture of novel real-world image libraries^30^.

### Synthetic data generation

Synthetic images of RDTs were mechanistically generated using 3D modelling software, Unity. This allowed control of all parameters of each image using C# scripts and features in Unity. Controlled parameters are detailed below and can be grouped into test specific and condition specific parameters. A detailed breakdown of example parameters and images that can be controlled is available in Supplementary Figure 1 and Supplementary Table 3.

Test specific parameters included test size, test result, control and test line sizes, opacities, appearances (solid and particle) and positions, test strip colour, simulations of liquid progressing into the ROI, appearance of randomised alphanumeric text above and to the right of the test window, appearance of outcome labelling text to the left-hand side of the test window, and appearance of symbols below the test window. A thin or thick solid test line was used to replicate homogeneous control and test line results, with intensity set between 0.5 to 100% opacity, while multiple overlayed particle systems fixed to two dimensions of varying sizes, number and area were used to replicate non-homogenous real-world test lines. Condition specific parameters included lighting, camera yaw, position of the camera by translating randomly in three dimensions, camera layer culling masks, camera field of view, camera aspect ratio, and output image size. Most parameters were governed by probabilistic mechanisms to create a wide variety of image properties. This probabilistic variation across multiple parameters results in a virtually infinite number of unique images. The image creation parameters were configured differently depending on the desired training dataset.

Images for training the segmentation model were created in pairs. In one image, the ROI (test window) can be assigned a clear block colour receiving no shadows (green shading), and the full cassette assigned a different clear block colour receiving no shadows (purple shading) (Fig. 1a, Supplementary Figure 6 and 7). A python script annotated the location of the ROI in each image pair. 4871 training images and annotated ROIs were used for training the segmentation model. 4000 positive and 4000 negative images were generated to train the HIV RDT image classification model (Supplementary Figure 8). 1000 positive and 1000 negative images were generated to train the COVID-19 RDT image classification model. All segmentation and classification synthetic images are available via the Data availability statement

The Supplementary Information and material available through the Code availability statement and Data availability statement provides all information required to produce synthetic images for a new RDT, including the 3D assets (and video outlining the creation of 3D assets), C# code and a detailed text guide.

### Image processing

Image processing consisted of several transformations aimed at standardising and improving data before training. Training images for the segmentation model were processed using various data augmentation techniques, including scaling images, mosaicking, random erasing, adjusting Hue, Saturation and Value (HSV), rotations, translations, and other data augmentation techniques (see Code availability statement for full training script). These were applied randomly through a predefined augmentation policy (RandAugment, https://docs.ultralytics.com/modes/train/).

For the HIV classification model, training images were subject to four image processing steps: resizing to 224 by 224 pixels preparing the image for the model input; blurring by applying a random intensity blur filter to soften image details; inverting by subtracting the value of each pixel from 1; and, finally, normalising by subtracting the mean pixel value and dividing by the standard deviation of all pixels in the image, closely following previous image processing to allow comparison^7^. Real-world images were subject to the same image processing excluding blurring. Blurring was applied only to synthetic training images to better approximate optical blur and noise characteristics of real-world image capture.

Training images for the COVID-19 classification model had slightly different processing steps to reduce computational demand. Training images were first segmented to the ROI using a segmentation model, resized to 224 by 224, normalised by dividing each pixel value by 255, blurred and inverted. For the real-world images, images were segmented, normalised, rescaled to emphasise pixel intensities above a threshold of 0.1, suppressing background noise and enhancing contrast in the test region, and finally inverted. As with the HIV classification model, blurring was applied only to synthetic training images. Inversion improved contrast between background strip and test region of positive results.

### Image labels

All synthetic images were labelled according to the virtual camera capturing the image from the mechanistic simulation environment. The C# script for generating image parameters, layer assignment and camera culling masks were checked before image generation to ensure correct labelling. Parameter extremes, such as highest light intensity and lowest test line intensity, were produced and visually checked to ensure the test outcome label was accurate.

The HIV RDT real-world image library was labelled by a panel of three RDT experts^7^. The COVID-19 RDT real-world image library was labelled as per previous publication^9^. All real-world RDT images were classified by ML into two outcome categories (positive/negative).

### Machine learning: training

For both classification models, a base model of MobileNetV2 was used with weights pre-trained on the ImageNet dataset, excluding the top layer. The model was prepared to receive images of 224 by 224 pixels with three channels corresponding to Red, Green, and Blue (RGB). After the base model, the output undergoes Global Average Pooling reducing the spatial dimensions to a single vector per image. This vector is processed through a fully connected layer consisting of 256 neurons with rectified linear unit (ReLU) activation. A single dropout layer (rate=0.5) was then applied before the output layer. For the HIV classification model, the final dense layer is a single neuron with a sigmoid activation function outputting a probability value. This is the same as previously described^7^ with two changes: the number of epochs was reduced from 50 to 10 in order to output high performance with faster training, and image preprocessing was updated to include the aforementioned blurring and inverting steps. Models were trained using GPU for comparison to previous work and training speed. For the COVID-19 classification model, training was conducted on a CPU demonstrating the technological accessibility of model development.

For the segmentation model, a nano YOLOv11 segmentation (yolo11n-seg.pt) base model was used, pre-trained on Ultralytics’ library^26,31^. A custom synthetic dataset defined in a YAML file, including synthetic images and annotations, was used for further training. The model was trained for 5 epochs on images resized to 640 by 640 pixels. Mixed precision training was used to balance memory usage and efficiency. All training scripts are either previously published^7^ or available via the Code availability statement. The trained model provided with the study documents is subject to the AGPL-3.0 license, an Open-Source Initiative approved open-source license, consistent with the licensing of the underlying YOLOv11 framework^26,31^.

The ClassifyAI and AutoAdaptPOC models were trained as previously described^7,9^. Briefly, ClassifyAI is a classification model built on MobileNetV2 pretrained on ImageNet and fine-tuned on 5,528 real-world images^7^. AutoAdaptPOC features automated segmentation using Mask R-CNN, an instance segmentation model pre-trained on the ImageNet1K dataset, combined with RDT kit and ROI dimensions, along with 60 real-world images to pre-train a segmentation model and 20-75 images to adapt to each new RDT kit. AutoAdaptPOC classifies images using a ResNet18 model pretrained via self-supervised learning on the ImageNet1K dataset and 383 real-world images and adapted using supervised contrastive learning on 10-14 labelled images^9^.

### Machine learning: best threshold

For the HIV RDT models, following training of the classification model on synthetic data, a best threshold was calculated by using 18% of the full dataset, the same proportion of data for validation as the benchmark ClassifyAI model trained on real-world data^7^. First, receiver operating characteristic curves (ROC) were constructed for the validation dataset, providing false positive rate (FPR (1 – specificity)) and true positive rate (TPR (sensitivity)) across many thresholds. The best threshold was determined by selecting the threshold which maximised the sum of TPR and 1 – FPR, maximising true positives and minimising false positives. The ratio of classes in the validation set was matched to the 1:8 positive to negative ratio in the full HIV RDT image library, as conducted for ClassifyAI. For the models trained on increasing real-world data (Fig. 2b), a 1:8 positive to negative ratio was also maintained in training datasets. For the first training dataset (N = 54, positive n = 5), the 18% validation dataset had 1 positive image and 8 negative images. For each for the increasing real-world image datasets this ratio is maintained.

For the COVID-19 RDTs, no real-world RDT images were used during training, validation, or best threshold calculation. Instead, a separate synthetic image library which the classification model was not trained on was used to calculate the best threshold by maximising the sum of TPR and 1 – FPR, as detailed above. This best threshold was then applied to all three COVID-19 RDTs.

### Machine learning: evaluation

For the HIV RDTs, evaluation subsets of real-world images were classified using a threshold derived from real-world data. This process was repeated across 10 random subsets and performance metrics - sensitivity, specificity and accuracy - are reported with error bars representing standard deviation from the mean, following previously described methods^7^. For the COVID-19 segmentation model, a random subset of 50 images from all positive and negative results were selected for each of the three COVID-19 RDTs and the ROIs were manually annotated. This manual annotation was used as the ground truth and compared against ML segmentation of each image to calculate the Intersection over Union (IoU), Dice coefficient and centroid errors (normalised to diagonal) for each RDT, with means and 95% confidence intervals reported. For the classification of COVID-19 RDTs, the complete real-world image libraries were classified based on the synthetically derived best threshold, and performance metrics are reported alongside 95% exact binomial (Clopper–Pearson) confidence intervals.

## Data availability

All synthetic images are available for immediate access through the following link: https://doi.org/10.5522/04/28351934. Source data for Fig. 2 is available in Supplementary Table 4 and Supplementary Table 5. The real-world image library used for evaluating the performance of HIV RDT classification models is available through the AHRI repository, as previously published: https://data.ahri.org/index.php/catalog/923#metadata-data_access. Source data for Fig. 3 is available in Supplementary Figure 9. The real-world image library used for evaluating the performance of the COVID-19 RDT segmentation and classification models is available through the following link, as previously published: https://figshare.com/projects/AutoAdapt/188457.

## Code availability

Custom code used in training and analysing the machine learning models presented in this study is available on GitHub at https://github.com/ElliottRogers/SynSight. The code was developed for Python 3.9. A detailed description of the code, including usage instructions and dependencies, is provided in several README files within the repository. This includes the machine learning models used in this study: HIV classification, COVID segmentation (based on the YOLOv11 framework and subject to the AGPL-3.0 license), and COVID classification. In addition, previously published, free and easy to access code used to train the ClassifyAI model and the classification models trained on increasing real-world HIV data (Fig. 2b) is available here: https://xip.uclb.com/product/classify_ai.

The code was primarily tested on macOS 14.6.1. Other operating systems may be compatible but were not extensively tested. The following hardware was used for training: HIV Classification, Apple M2 (GPU - Metal); COVID Segmentation, Apple M2 (GPU - Metal); COVID Classification: CPU (tested on Google Colab GPU).

## Clinical trial number

not applicable.

The authors declare no competing financial or non-financial interests.

## Acknowledgements

We would like to thank the authors, institutions and participants who contributed to collecting and providing access to real-world image libraries, as outlined in Turbé et al. and Arumugam et al. We acknowledge funding for this research: Medical Research Council for the PhD studentship (MR/W006774/1 to E.R); EPSRC Digital Health Hub for AMR (EP/X031276/1 to R.A.M); MS is an NIHR Research Professor (NIHR 301634); AHRI is supported by core funding from the Wellcome Trust (082384/Z/07/Z and 201433/Z/16/Z to KH and MS). For the purpose of open access, the author has applied a CC BY public copyright licence to any Author Accepted Manuscript version arising from this submission.

## Author Contributions

ER, RAM, MS, VT, KH contributed to the conceptualisation of the manuscript. RAM and MS supervised the study. All authors – ER, VT, DG, CH, RAM, KH, MS - read and commented on early drafts, and approved the final manuscript. KH was principal investigator of the original study that produced the real-world HIV RDT images.

