## Supplemental Information for "Synthetic Data to Lower Barriers Towards Equitable Artificial Intelligence in Rapid Diagnostic Test Interpretation"

**Contents**

A. **Supplementary Tables**
B. **Supplementary Figures**
A. **Supplementary Tables**

**Supplementary Table 1. Performance metrics of one segmentation model applied to three different RDTs.** A subset of 50 real-world images were manually annotated for ground truth. Table columns show the region of interest (ROI) detection rate, and, calculated against the ground truth and reported as mean and 95% confidence intervals, the Intersection over Union (IoU), Dice coefficient and centroid errors (normalised to diagonal).

| Test | ROI detected | IoU (95% CI) | Dice (95% CI) | Centroid error (95% CI) |
| --- | --- | --- | --- | --- |
| DeepBlue (n=50) | 100% | 0.855 (0.839, 0.871) | 0.921 (0.911, 0.930) | 0.002 (0.002, 0.003) |
| Jinwofu (n=50) | 100% | 0.832 (0.794, 0.869) | 0.899 (0.861, 0.937) | 0.003 (0.002, 0.003) |
| EcoTest (n=50) | 100% | 0.833 (0.814, 0.851) | 0.907 (0.896, 0.919) | 0.004 (0.004, 0.005) |

**Supplementary Table 2. Comparison of real-world training data use for each of the different SynSight models, across HIV classification, COVID-19 segmentation and COVID-19 classification.**

| **Model** | **Real-world data used** | | |
| --- | --- | --- | --- |
|  | **Training** | **Threshold** | **Evaluation** |
| HIV classification | No | Yes | Yes |
| COVID-19 segmentation | No | N/A | Yes |
| COVID-19 classification | No | No | Yes |

**Supplementary Table 3. False negatives and false positives identified by SynSight, classified by reason for error, across the three COVID-19 RDT image libraries (DeepBlue, N=283; Jinwofu, N=200; and EcoTest, N=610).** False negatives (FN) – ML negative, ground truth positive – are challenging to overturn due to limitations in result image capture. All FN results were ML errors due to weak test lines (8), dark image (1), poor region of interest segmentation (1), and errors that couldn’t be characterised (11). False positive (FP) – ML positive, ground truth negative – where ML was correct (4) could be resolved as a weak test line was visible in result images. FP where ML was incorrect were due to artefacts near the test line (2), streaking (1), and errors that couldn’t be characterised (1).

| **False negatives** (SynSight negative, ground truth positive) | | |  |
| --- | --- | --- | --- |
| **RDT** | **n** | **Reason** | |
| DeepBlue | 0 | - | |
| Jinwofu | 2 | Dark image (1), poor segmentation (1) | |
| EcoTest | 19 | Weak test line (8), not characterised (11) | |
| **False positives** (SynSight positive, ground truth negative) | | |  |
| **RDT** | **n** | **Ground truth error, ML correct** | **ML error** |
| DeepBlue | 2 | Weak test line (1) | Streaking (1) |
| Jinwofu | 4 | Weak test line (3) | Not characterised (1) |
| EcoTest | 2 | — | Artefact near test line (2) |

**Supplementary Table 4. Parameters of synthetic image generation that can be controlled through mechanistic simulation.**

| **Domain** | **Description** | **Details** |
| --- | --- | --- |
| Test geometry | Test dimensions | Size and shape of cassette or strip |
|  | Test details | Printed labels (eg. IgG, IgM, alphanumeric patient id) |
|  | Spatial position (x, y, z) | Position of test in image |
|  | Orientation (roll, pitch, yaw) | Rotation of test in image |
| Camera | Camera position (x, y, z) | Distance between camera and test |
|  | Camera orientation (roll, pitch, yaw) | Angle between camera and test |
|  | Scene layering | Visible layers or objects in camera view |
| Test line properties | Intensity | Opacity or contrast of test line |
|  | Morphology | Homo/heterogenous test line, thickness, particle distribution |
|  | Presence | Determines test result (positive, negative, invalid, IgG positive etc) |
| Scene | Background | Imaging surface (eg. package insert, wooden table) |
|  | Occlusion | Obstruction of full test |
| Lighting | Intensity | Bright or dark imaging |
|  | Colour | Colour of lighting |
|  | Angle | Lighting angle determining shadows |
|  | Shadow strength | Strength and disparity of shadows |
| Class distribution | Label frequency | Proportion of classes in dataset |
|  | Rare cases | Presence of underrepresented classes |
|  | Mask | Image label or image mask pairing |

**Supplementary Table 5. Sensitivity and specificity of 10-fold validation of machine learning models trained on increasing real-world images reported with mean ± standard deviation (n=54 to n=6221).**

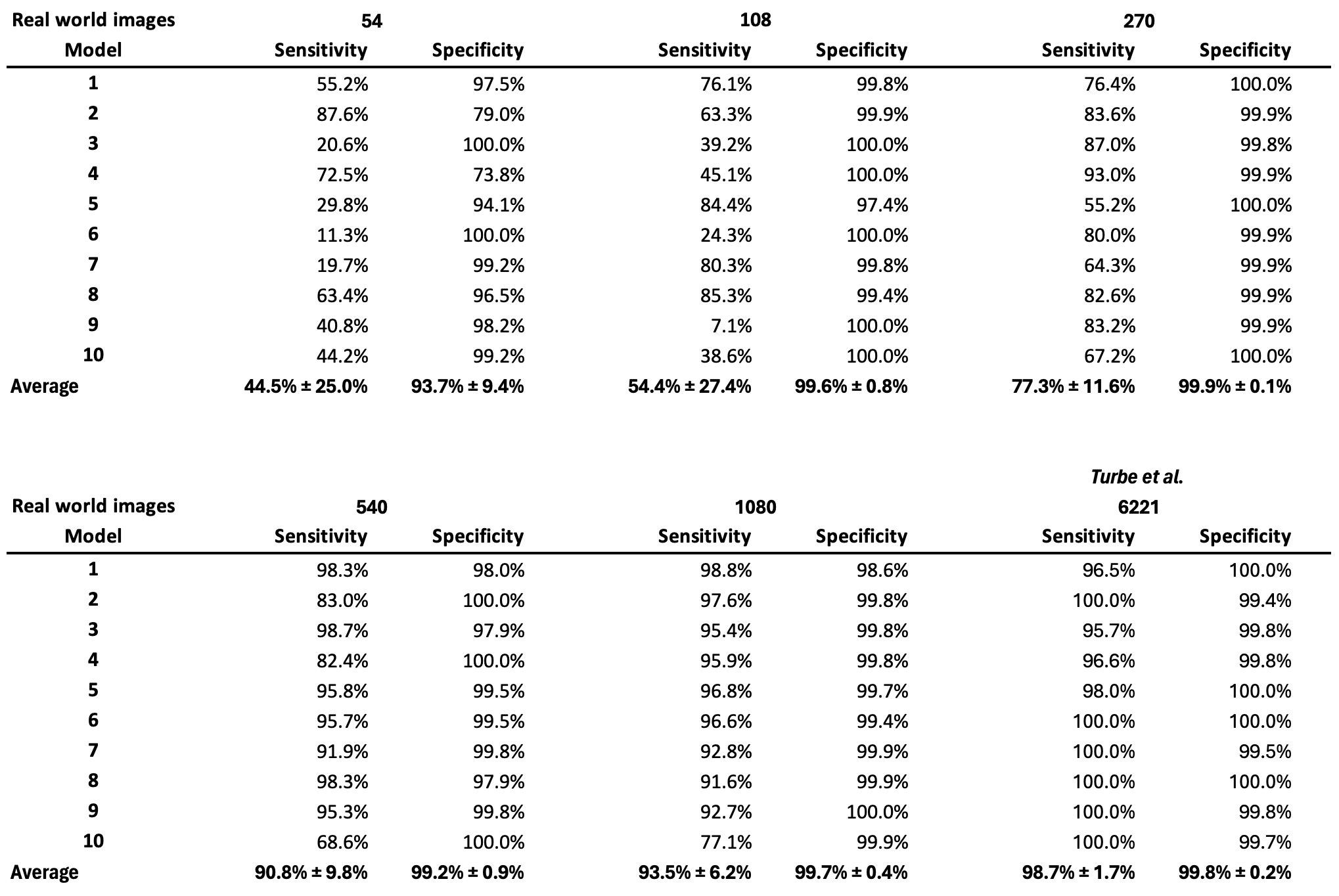

**Supplementary Table 6. Train on synthetic, test on real (TSTR) performance of our Machine Learning model compared to train on real, test on real (TRTR) performance of a model trained on 6,221 (*Turbé et al)*^1^ and 270 real-world images reported with mean ± standard deviation**.

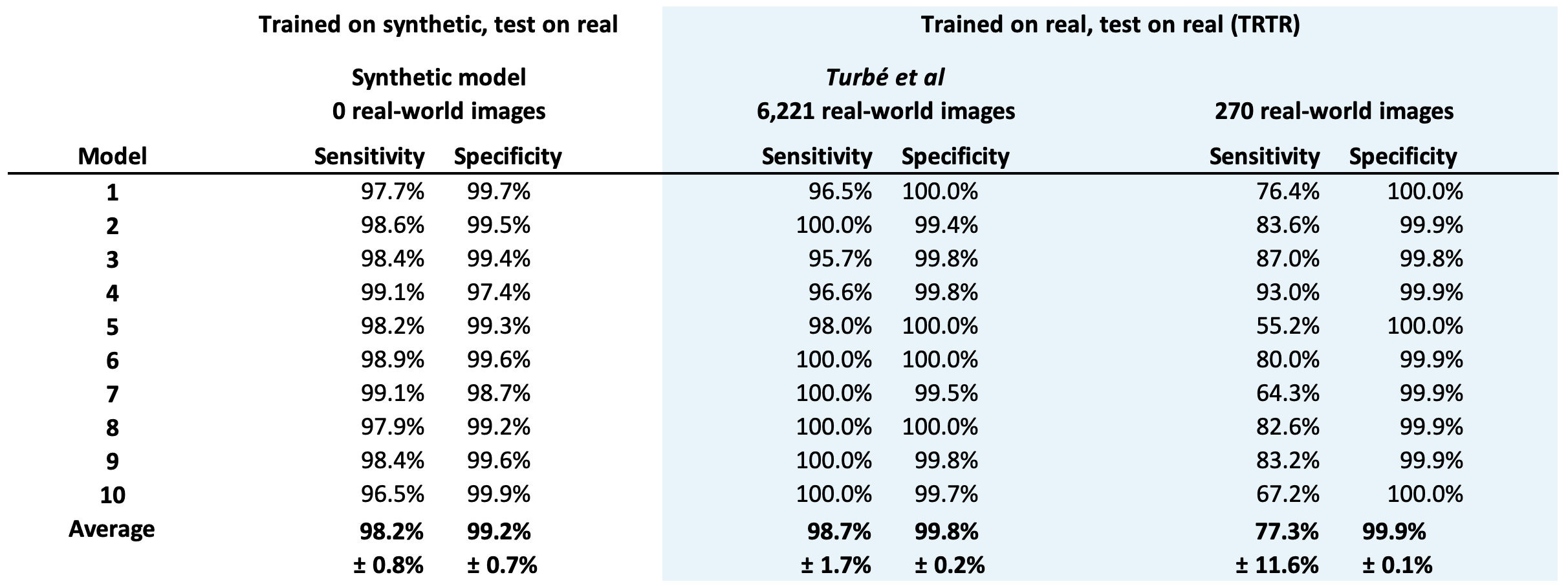

B. **Supplementary Figures**

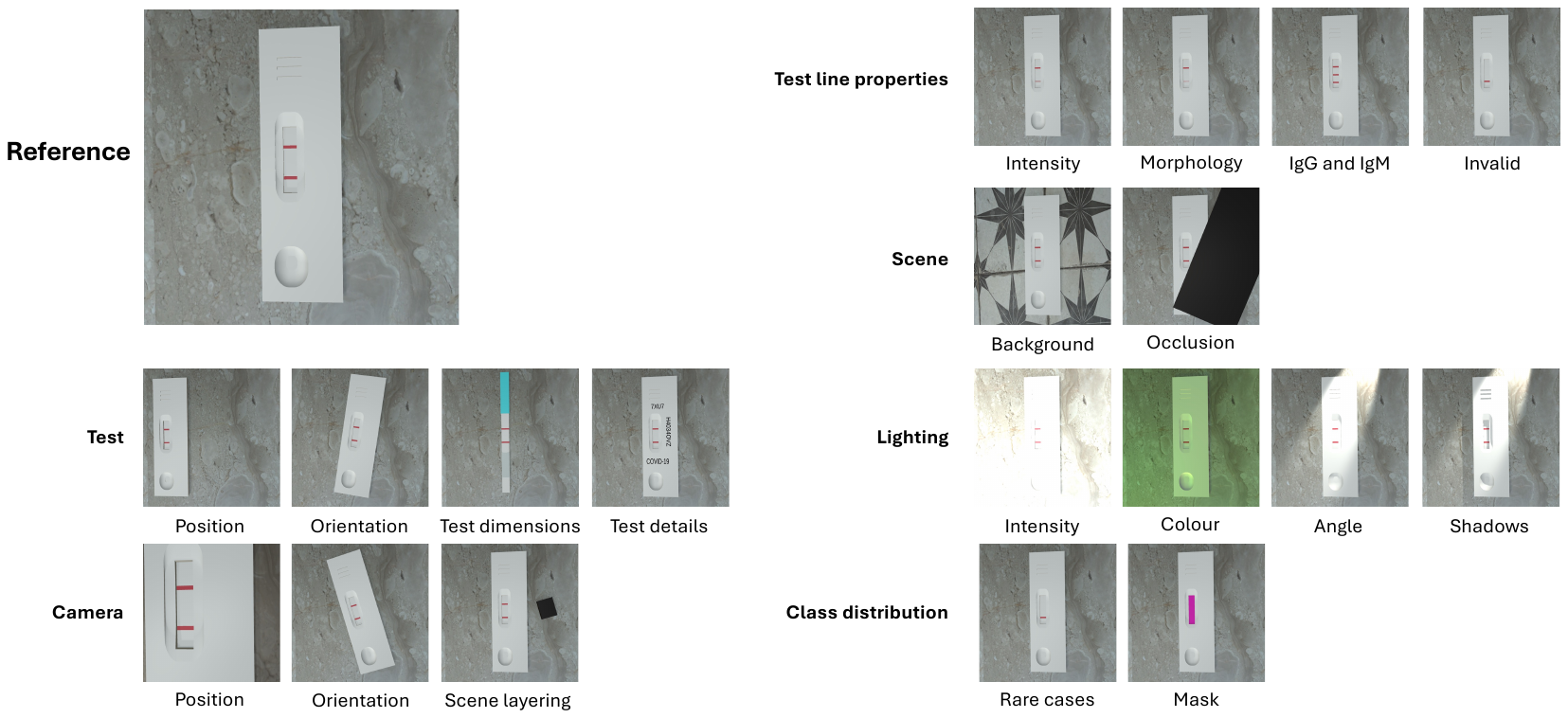

**Supplementary Figure 1. Synthetic images generated by changing image parameters alongside a reference image.** Example parameters that are controlled in 3D modelling software Unity relate to test (position, orientation, dimensions e.g. cassette and strip, details e.g. labelling); camera (position, orientation, scene layering); test line properties (intensity, morphology, test results e.g. positive or IgG and IgM or invalid); scene (background variation, occlusion of test); lighting (intensity, colour, angle, shadow strength); and class distribution (rare cases, and masking).

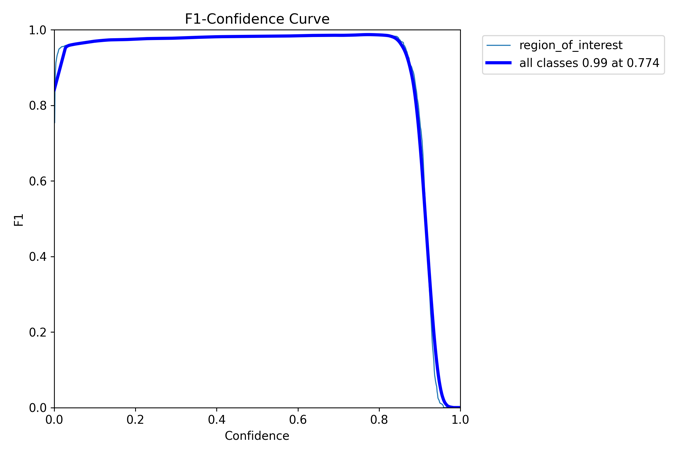

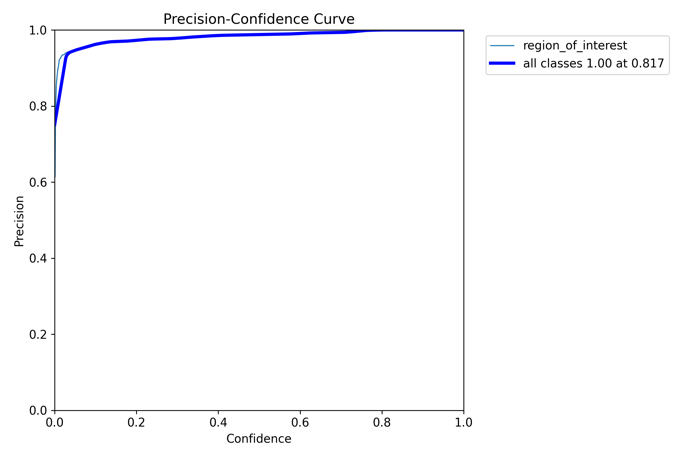

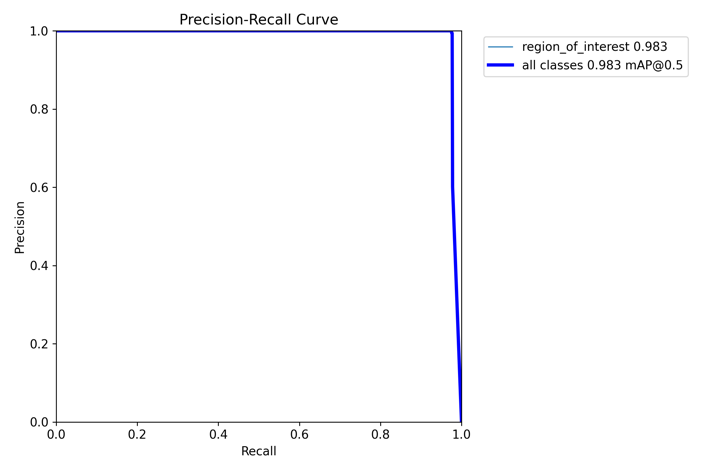

**Supplementary Figure 2.** Box F1-Confidence, Precision-Confidence, and Precision-Recall plots for the COVID-19 segmentation model.

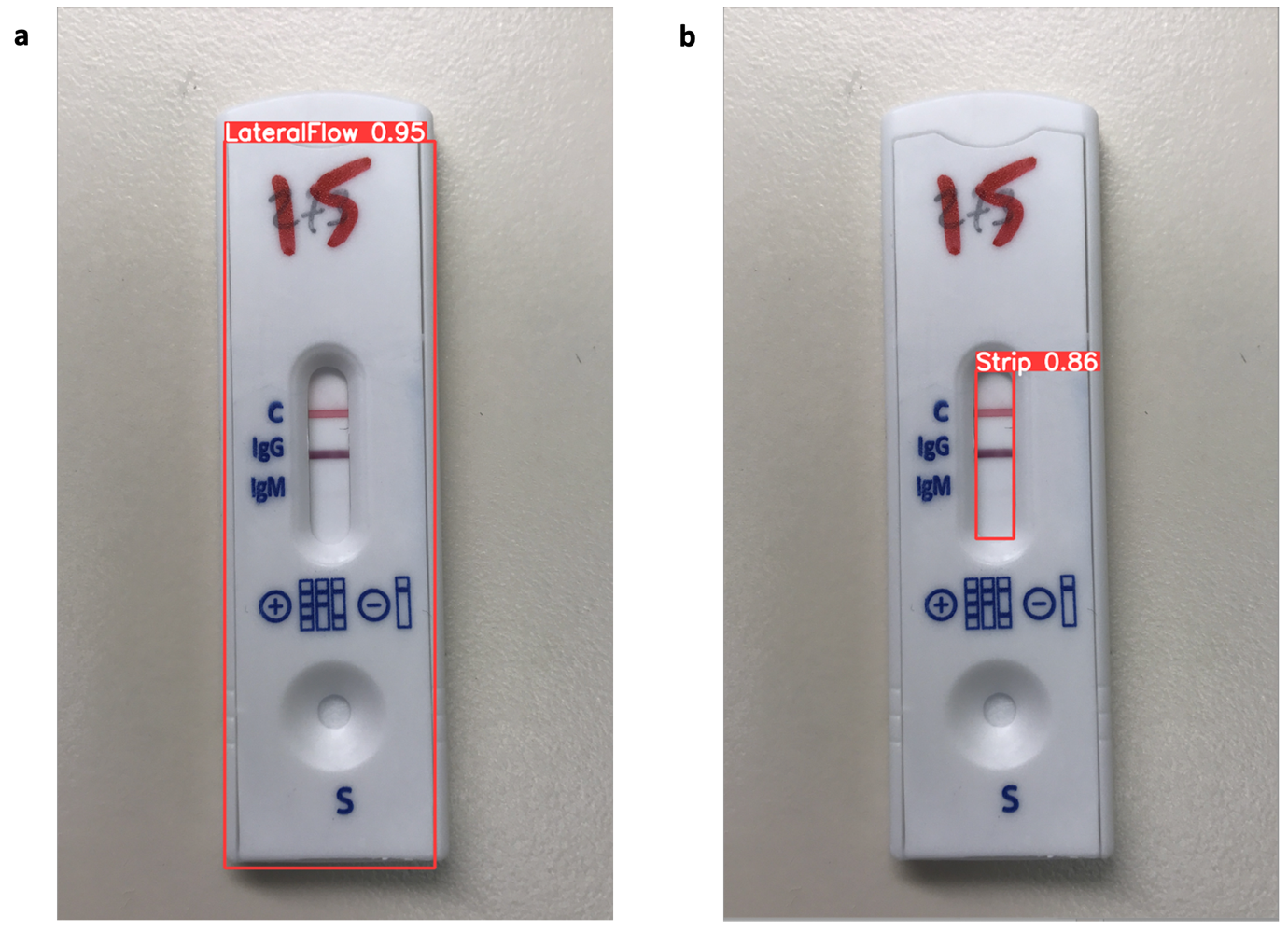

**Supplementary Figure 3. Augmented Reality (AR) overlays of object detection on COVID-19 RDTs.** The red rectangles indicate object detection of a lateral flow (a) and strip, or test window (b). These models could aid users in capturing high quality and standardised images during image capture. Real-world images collected by *Arumugam et al^2^*.

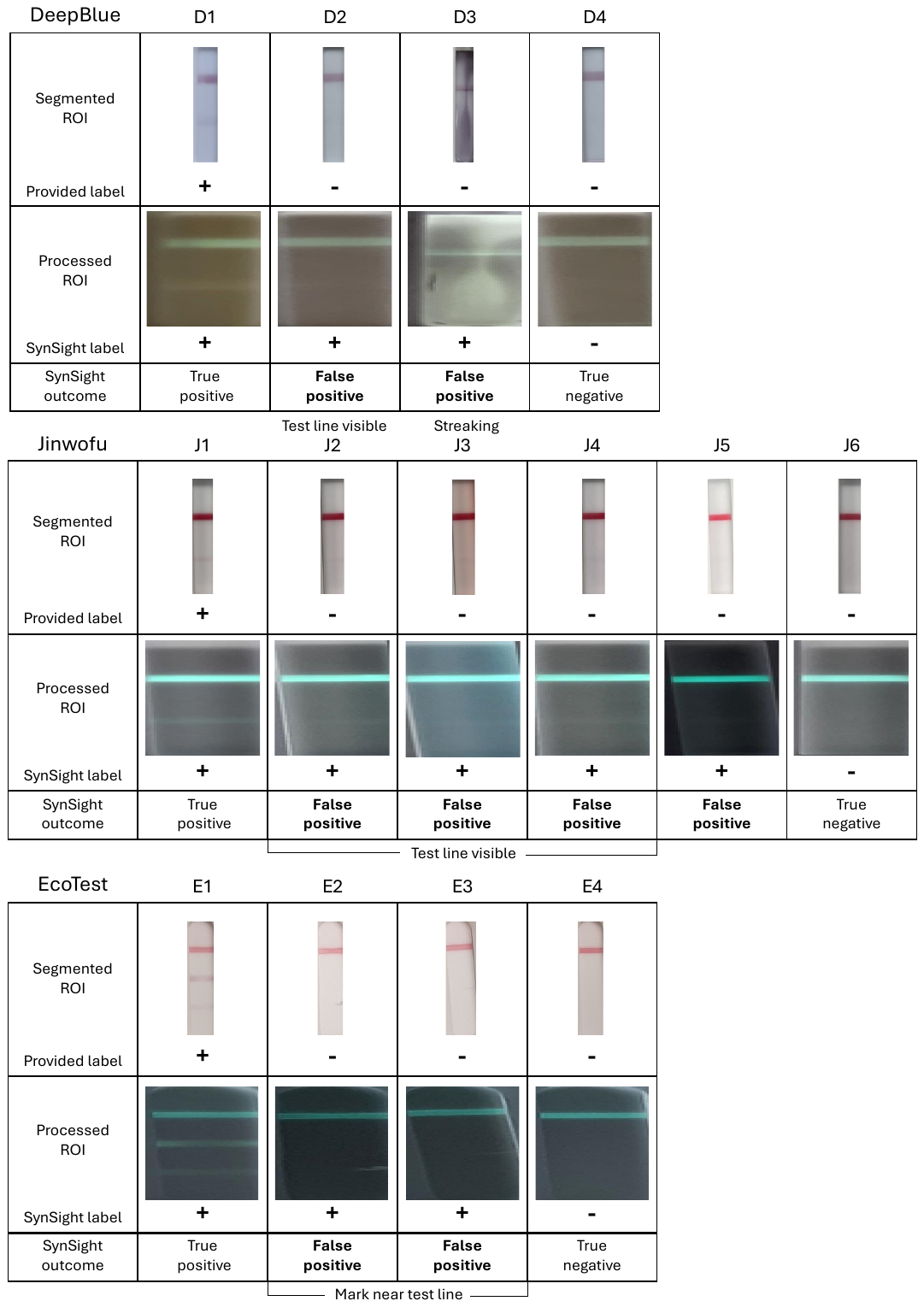

**Supplementary Figure 4. False positive results (SynSight positive and provided ground truth negative) for each of the three evaluated COVID-19 RDTs.** The two DeepBlue RDT false positives (D2, D3), four Jinwofu RDT false positives (J2-5), and two EcoTest RDT false positives (E2, E3) are presented alongside true positives (D1, J1, E1) and true negatives (D4, J6, E4). A positive test line is visible in four false positives (DeepBlue D2; Jinwofu J2-4), suggesting incorrect ground truth labelling. One DeepBlue RDT (D3) has streaking (nanoparticle distribution throughout the region of interest (ROI)), and two EcoTest RDTs (E2, E3) have marks near the test line location, likely resulting in the false positive result. Real-world images collected by *Arumugam et al^2^*.

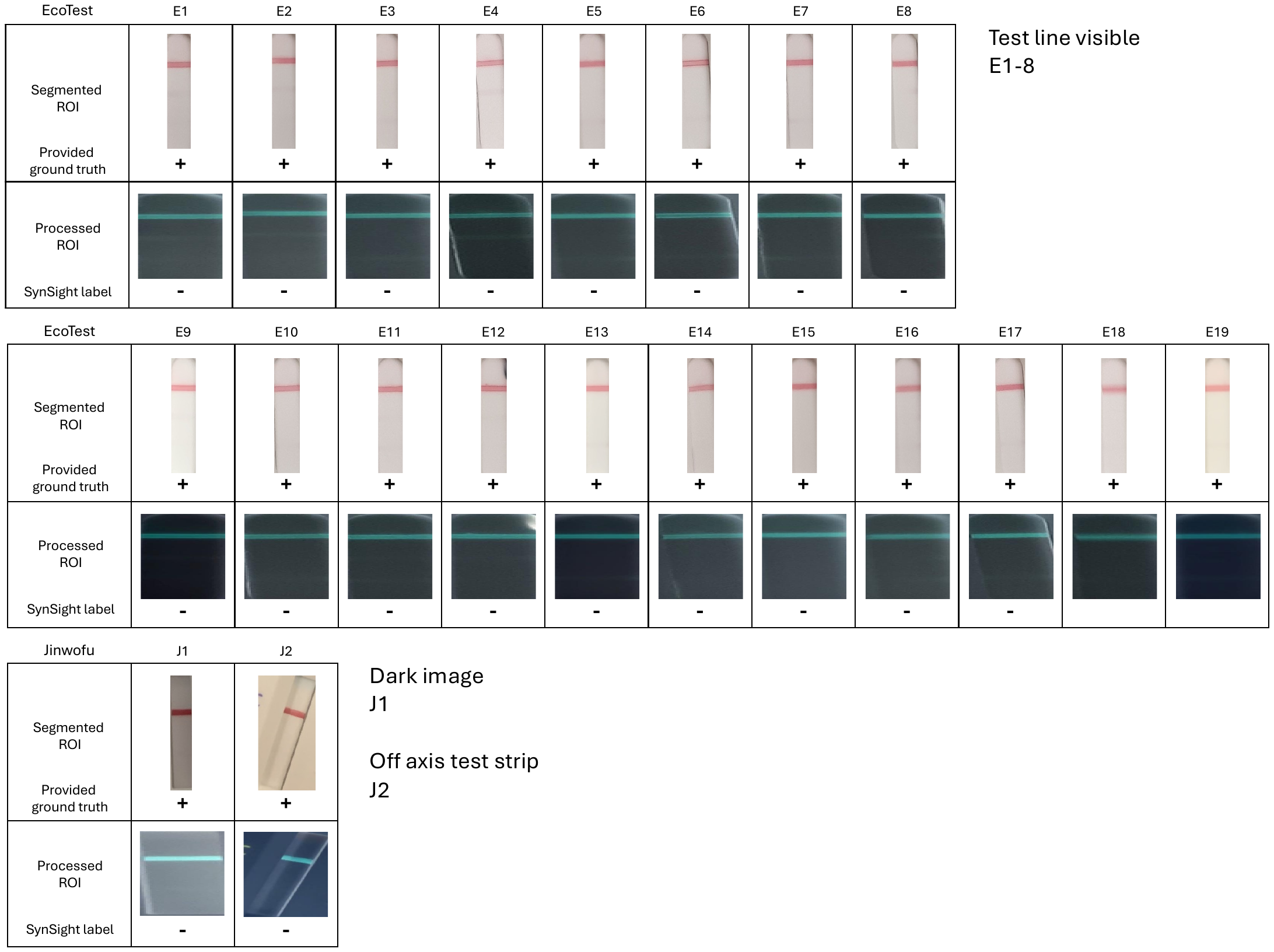

**Supplementary Figure 5. False negative results (SynSight negative and provided ground truth positive) of EcoTest (n=19) and Jinwofu (n=2) COVID-19 RDTs.** No false negative EcoTest RDT had strong test lines, while 8/19 had weak test lines. One false negative Jinwofu RDTs has dark shadows (J1) and one is captured off axis leading to poor segmentation (J2). Real-world images collected by *Arumugam et al^2^*.

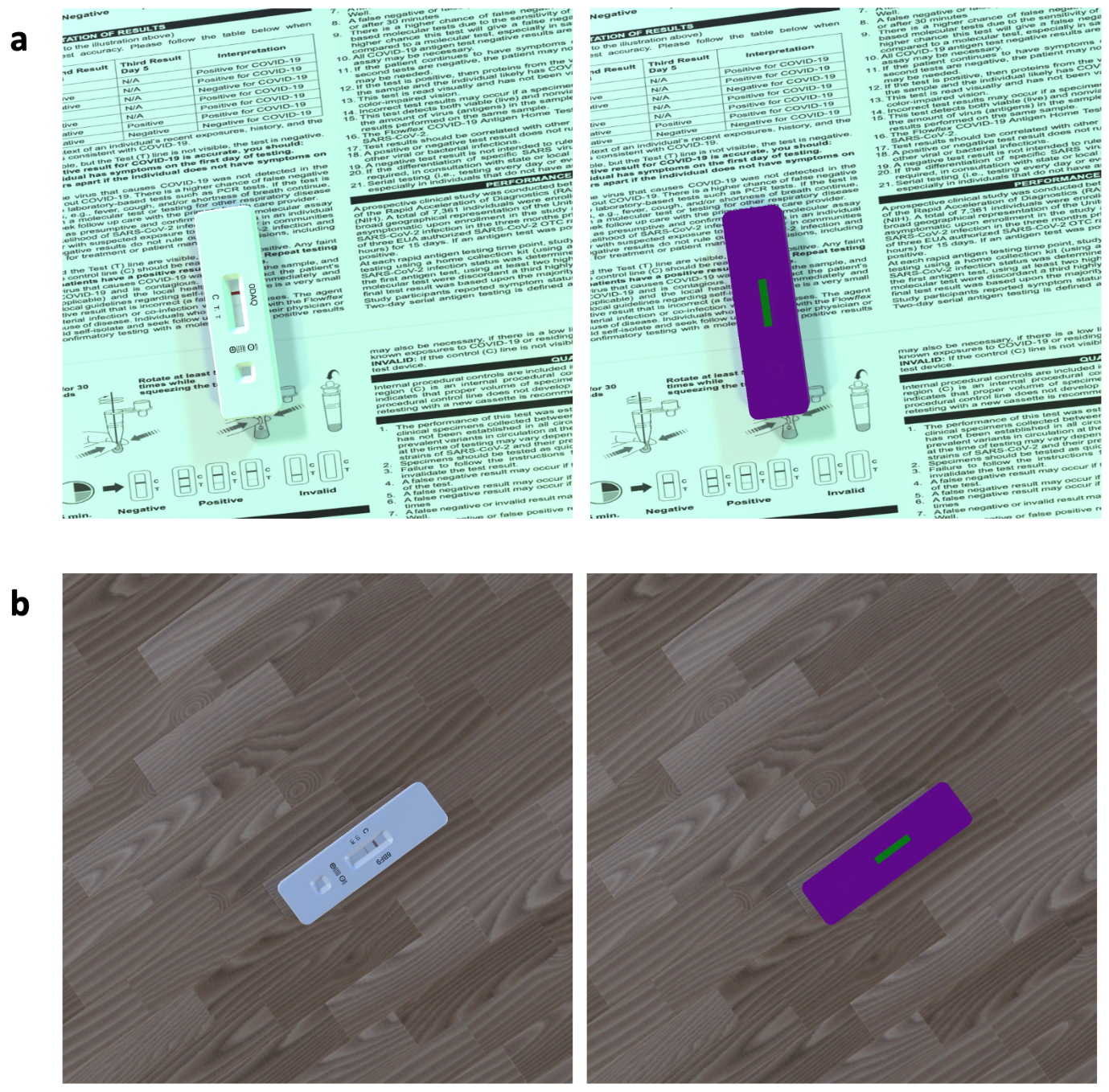

**Supplementary Figure 6. Close up comparison of two real-world images used during training of the segmentation model.** a, and b, show two examples of RDT images generated using 3D modelling software Unity as part of the synthetic dataset used to train a segmentation model with YOLOv11. The left hand side images show the normal synthetic image of the lateral flow with varying background, light colour, light intensity, labelling, lateral flow orientation and zoom. The right-hand side images show duplicate images where the materials of the lateral flow cassette and test window are replaced materials receiving no shadows or lighting from the environment. This allows masks applied through python scripts to identify the lateral flow (purple) and the test window (green). Coordinates of these areas can then be combined with the left-hand side image to generate the training dataset needed, as outlined by YOLOv11 documentation (<https://docs.ultralytics.com/tasks/classify/#models>).

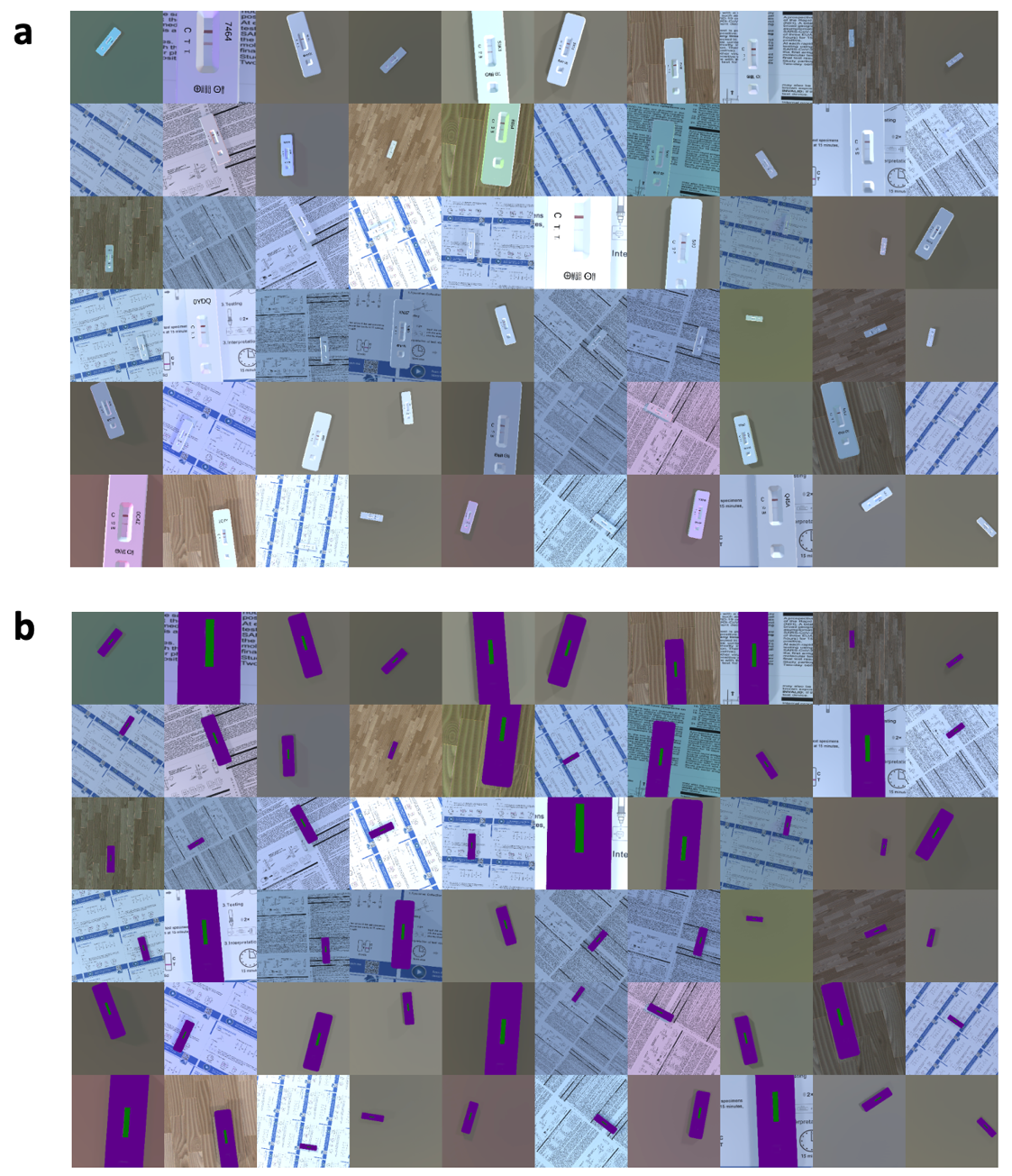

**Supplementary Figure 7. Examples of synthetic images used to train segmentation models. a,** 60 example synthetic images generated using 3D modelling software Unity showing positive results used during training classification model. **b,** The corresponding 60 example real-world images showing positive results with a purple material replacing the lateral flow and a green material replacing the test window.

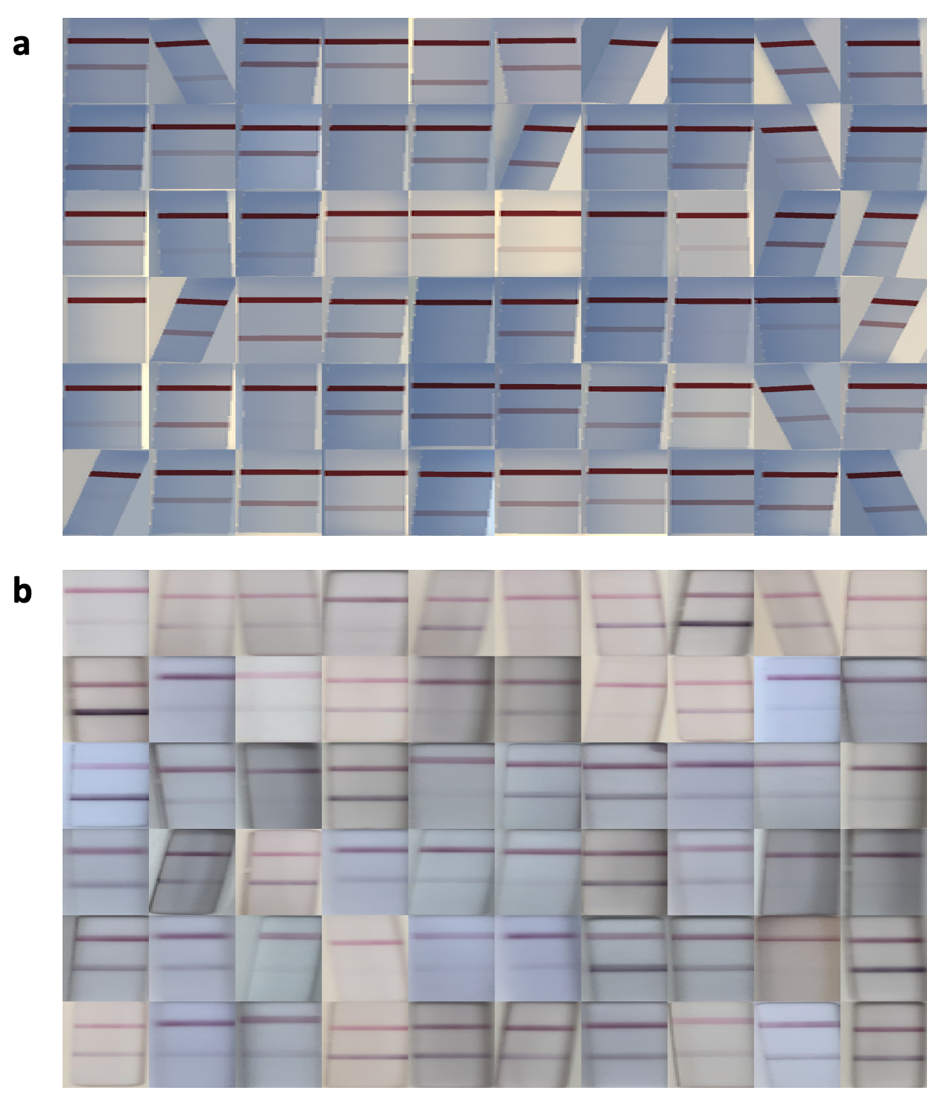

**Supplementary Figure 8. Examples of images used during training and classification of COVID-19 RDTs. a**, 60 example synthetic images generated using 3D modelling software Unity showing positive results used during training of the COVID-19 RDT classification model. **b**, 60 example real-world images collected by *Arumugam et al^2^* showing segmented positive results of the DeepBlue COVID-19 Antibody RDT.

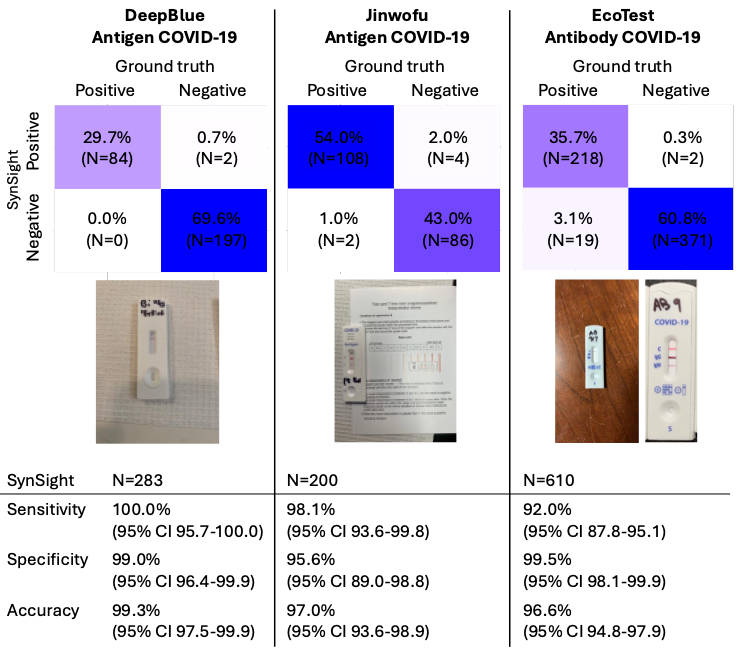

**Supplementary Figure 9. Example images and confusion matrices of the full Synthetic Machine Learning (SynSight) pipeline for three COVID-19 RDTs**. High accuracy is seen across all three RDTs: DeepBlue COVID-19 Antigen (TSTR: 99.3%, 283 images classified); Jinwofu COVID-19 Antigen (TSTR: 97.0% accuracy, 200 images classified); and EcoTest COVID-19 Antibody (TSTR: 96.6%, 610 images classified). The Jinwofu COVID-19 Antigen RDT image illustrates a printout interpretation support document which is part of the segmented image, which does not affect segmentation of the ROI or classification of the result. The EcoTest images illustrate the variability in RDT size within each image. Real-world images collected by *Arumugam et al^2^*. All metrics are reported with 95% exact binomial (Clopper–Pearson) confidence intervals (CI).
